# Effects of genomic recombination on SARS-CoV-2 evolution and the growth of the recombinant variant XFG in Germany

**DOI:** 10.1101/2025.11.17.25339653

**Authors:** Buqing Yi

## Abstract

In recent several years, multiple predominant SARS-CoV-2 variants that spread worldwide were derived from genomic recombination between SARS-CoV-2 lineages with diversified genetic backgrounds. However, the current understanding about the effects of recombination on SARS-CoV-2 evolution and functional aspects is limited.

In this study, to achieve one overview regarding the evolution of SARS-CoV-2 recombinant variants, phylogenetic analyses have been performed to evaluate the divergence of representative Omicron recombinant variants from the predicted original Omicron lineage as well as the phylogenetic distances among these variants. As one example of predominant recombinant variants, we also investigated the growth of XFG in Germany by means of virus genomic epidemiology approaches.

The results of the evolutionary relationship analyses indicate that recombination between evolutionarily distant lineages or closely related lineages can both drive SARS-CoV-2 evolution, and has the potential of resulting in a novel predominant strain, indicating that recombination plays one important role in SARS-CoV-2 evolution. Furthermore, this study provided information about the growth of XFG in Germany, and revealed a clear relative growth advantage of XFG over co-circulating lineages, such as LP.8.1 and NB.1.8.1. The information acquired from these investigations underlines the need for continuous efforts to detect recombination events and track recombinant variants, which is important for evaluating the long-term effects of recombination on SARS-CoV-2 evolution and of significance for public health.

## 1. Introduction

SARS-CoV-2 recombinants brought challenges for detection, characterization, and evaluation of the phenotype of the virus ^1,2^. Nevertheless, it is essential to track recombinant genomes because they contain a unique combination of mutations and have possible impacts on pathogenicity or other functional aspects of the virus, which is critical in the context of public health. Novel strains derived from recombination between different SARS-CoV-2 variant genomes have the potential of evading host immunity gained from previous infections or vaccination, which may possibly affect the spread of the strains ^3,4^. Furthermore, recombination between different coronaviruses was assumed to play a role in the origin of SARS-CoV-2 ^5-8^, indicating the importance of monitoring recombinant viruses in protecting public health.

It is known that coronaviruses could produce many kinds of recombination products during natural infection, including recombinant genomes ^9^. Recombinant SARS-CoV-2 genomes arise from recombination of two or more different SARS-CoV-2 lineages and often appear following the co-circulation of multiple lineages at high prevalence, which may lead to co-infection of specific individuals. Co-infection provides the circumstances during which chimeric genotypes could emerge, typically through template switching or homologous recombination ^10^. Several studies have reported discovery of recombination events within a single patient co-infected with co-circulating lineages ^11,12^. However, there is also genomic evidence suggesting that recombination between non-co-circulating viruses may occur within long-term-infected individuals, wherein the original lineage recombines with a more recent lineage from a subsequent infection ^12,13^.

The recombinant lineage XA, derived from recombination between B.1.1.7 (the Alpha variant) and B.1.177 genomes, is the first identified SARS-CoV-2 recombinant lineage in the Pango nomenclature system ^14,15^. XA emerged in Europe in early 2021 following the local co-circulation of B.1.1.7 and B.1.177 ^16^. It is noteworthy that prior to the designation of XA, in the first year of the pandemic, certain SARS-CoV-2 recombinants already emerged and circulated at low levels ^17^. Following the designation of XA, more and more recombinant lineages have been identified ^18^. The latest SARS-CoV-2 recombinant is represented by the variant XFG (nicknamed “Stratus” or “Frankenstein”), which was first detected in January 2025 and designated as Variant under Monitoring (VUM) by the WHO in the middle of 2025. The fast increase of XFG-caused infections suggests the necessity of acquiring detailed information about the spread of this recombinant variant.

During the pandemic, many designated recombinant SARS-CoV-2 lineages and a few newly emerging variants were identified through the genomic surveillance efforts in Germany ^19-21^, and SARS-CoV-2 genomic data has been further produced after the pandemic in view of public health protection as well as research purpose ^22-26^. The objective of this study is to evaluate the effects of recombination on SARS-CoV-2 evolution through phylogenetic analyses, and also to investigate the emergence and spread of the recombinant variant XFG in Germany via genomic epidemiology approaches.

## 2. Methods

### 2.1 Phylogenetic analysis of SARS-CoV-2

For phylogenetic analysis, we used samples collected worldwide and uploaded to GISAID ^27^. Only samples fulfilling these quality check criteria on GISAID were included in the analysis: 1. With a complete sequence (>29,000nt) and less than 5% Ns; 2. With complete sample collection dates. And the standard quality assessment parameters utilized in NextClade (https://clades.nextstrain.org) ^28^ as described in 2.2, were applied as well.

Phylogenetic analyses were carried out to investigate evolutionary relationship with a custom build of the SARS-CoV-2 NextStrain build (https://github.com/nextstrain/ncov) ^19-21,29^. The pipeline includes several Python scripts that manage the analysis workflow. Briefly, it allows for the filtering of genomes, the alignment of genomes in NextClade (https://clades.nextstrain.org) ^28^, phylogenetic tree inference in IQ-Tree ^30-32^, tree dating ^33^ and ancestral state construction and annotation. In this investigation about the evolutionary relationship among representative recombinant lineages, the phylogenetic tree was rooted with “least-squares” methods ^34^ to make the phylogeny less affected by square errors of the branch lengths and therefore more accurately reflect sample-to-sample relationship. For each recombinant lineage, around ten samples were randomly chosen from the sequences that passed the quality check and used for the analysis.

### 2.2 Monitoring the spread of SARS-CoV-2 in Germany through genomic epidemiology investigation

We combined SARS-CoV-2 sequences generated from samples collected in Germany (all the sequences were shared on GISAID ^27^) to build up genome sequence data set for epidemiology investigation. We performed quality check and filtered out low-quality sequences that met any of the following criteria: 1) sequences with less than 90% genome coverage; 2) genomes with too many private mutations (defined as having >24 mutations relative to the closest sequence in the reference tree); 3) genomes with more than ten ambiguous bases; and 4) genomes with mutation clusters, defined as 6 or more private differences within a 100-nucleotide window. These are the standard quality assessment parameters utilized in NextClade (https://clades.nextstrain.org) ^28^. In the current study, between January 2025 and July 2025, more than 1,600 sequences from Germany were used in the genomic epidemiology analyses. For lineage identification, we used the dynamic lineage classification method through the Phylogenetic Assignment of Named Global Outbreak Lineages (PANGOLIN) software suite (https://github.com/hCoV-2019/pangolin) ^14^.

### 2.3 Relative growth advantage

We also analyzed relative growth advantage with SARS-CoV-2 sequences generated from samples collected in Germany from the defined time duration. A logistic regression model was used to estimate the relative growth advantage of certain variant compared to co-circulating variants as previously reported ^35-38^. The model assumes that the increase or decrease of the proportion of a variant follows a logistic function, which is fit to the data by optimizing the maximum likelihood to obtain the logistic growth rate in units per day. Based on that, an estimate of the growth advantage per generation is obtained (assuming the growth advantage arising from a combination of intrinsic transmission advantage, immune evasion, and a prolonged infectious period ^39^, and the relative growth advantage per week (in percentage; 0% means equal growth) is reported. The relative growth advantage estimate reflects the advantage compared to co-circulating variants in the selected region and time frame. The analyses were primarily performed with RStudio v1.3.1093 with multiple R software, e.g. tidyverse, ggplot ^40-46^.

## 3. Results

### 3.1 Recombination may drive the evolution of SARS-CoV-2

Since the designation of the first recombinant lineage XA in 2021, in the past several years, various recombinant variants have been identified, and some of them were defined as Variant of Interest (VOI) or Variant under Monitoring (VUM), which generally grow fast and circulate worldwide. The genetic backgrounds of the recombinant strains are diversified, and one overview analysis of the evolutionary relationship among the recombinant strains is lacking. In this study, we investigated the effects of recombination on SARS-CoV-2 evolution with phylogenetic analyses and also examined the evolutionary relationship among multiple representative SARS-CoV-2 recombinant variants.

In Table 1, we list representative SARS-CoV-2 recombinant variants and their known source lineages which were identified previously (the identification refers to the information provided in the Pango nomenclature system ^14,15^ and the relevant publications). For the clarity of the information, a few VOIs/VUMs as decedents of the recombinant variants are listed as well, such as XBB.1.5 and NB.1.8.1, with their ancestor lineage being indicated. XA, XB, XC, XD, XE, XF are among the earliest designated recombinant lineages. XA, XB and XC were produced in the pre-Omicron time ^47^; while both XD and XF were from recombination between Delta and Omicron ^51^. XE was from recombination between two Omicron lineages: BA.1 and BA.2. There are also several recombinants of BA.2 and BA.5, such as XAZ (from BA.5 and BA.2.5), which only showed limited circulation. Multiple predominant lineages in the past several years were derived from recombination, such as XBB.1.5, XEC, NB.1.8.1 and XFG.

**Table 1:** List of representative SARS-CoV-2 recombinant variants and genetic background.

| Recombinant Variant | Source Lineages |  |
| --- | --- | --- |
| **XA**<sup>16</sup> | B.1.1.7 (Alpha) | B.1.177 |
| **XB**<sup>47</sup> | B.1.631 | B.1.634 |
| **XC**<sup>48</sup> | AY.29 (Delta subvariant) | B.1.1.7 (Alpha) |
| **XD** | AY.4.2.2 (Delta subvariant) | BA.1 (Omicron lineage) |
| **XE** | BA.1 (Omicron lineage) | BA.2 (Omicron lineage) |
| **XF** | AY.4 (Delta subvariant) | BA.1 (Omicron lineage) |
| **XAZ** | BA.2.5 (BA.2 subvariant) | BA.5 (Omicron lineage) |
| **XBB** | BJ.1 (BA.2 subvariant) | BM.1.1.1 (BA.2 subvariant) |
| **XBB.1.5**<sup>49</sup> | XBB.1 | - |
| **XDE** | GW.5.1 (XBB subvariant) | FL.13.4 (XBB subvariant) |
| **XDR**<sup>50</sup> | JD.1.1 (XBB subvariant) | JN.1.1 (BA.2.86 subvariant) |
| **XDV** | XDE | JN.1 (BA.2.86 subvariant) |
| **XEC** | KS.1.1 (JN.1 subvariant) | KP.3.3 (JN.1 subvariant) |
| **XFG** | LF.7 (JN.1 subvariant) | LP.8.1.2 (JN.1 subvariant) |
| **NB.1.8.1** | XDV.1.5.1 | - |

In the phylogenetic analyses, we focus on representative Omicron recombinant variants, in particular the VOCs/VOIs/VUMs that got spread worldwide. In Figure 1, the values of “Divergence” (calculated through the relevant algorithm applied in Nextstrain) of each variant from the predicted common ancestor of BA.1 and BA.2 (the earliest Omicron lineages) are displayed. Based on evolutionary relationship, most current circulating Omicron recombinant variants were derived from BA.2. Before the emergence of XBB* (XBB and its subvariants), although various recombinant variants were identified, most of them showed limited circulation. XBB.1.5 is the first recombinant lineage that was independently identified as VOI. Thereafter, a few other XBB subvariants were also defined as VOI or VUM, such as XBB.1.9.1 and XBB.1.16 ^52^. The ancestor lineage XBB was produced by recombination between two BA.2 subvariants: BM.1.1.1 and BJ.1. The XBB* lineage family is evolutionarily more closely associated with BJ.1 (Figure 1).

**Figure 1:**
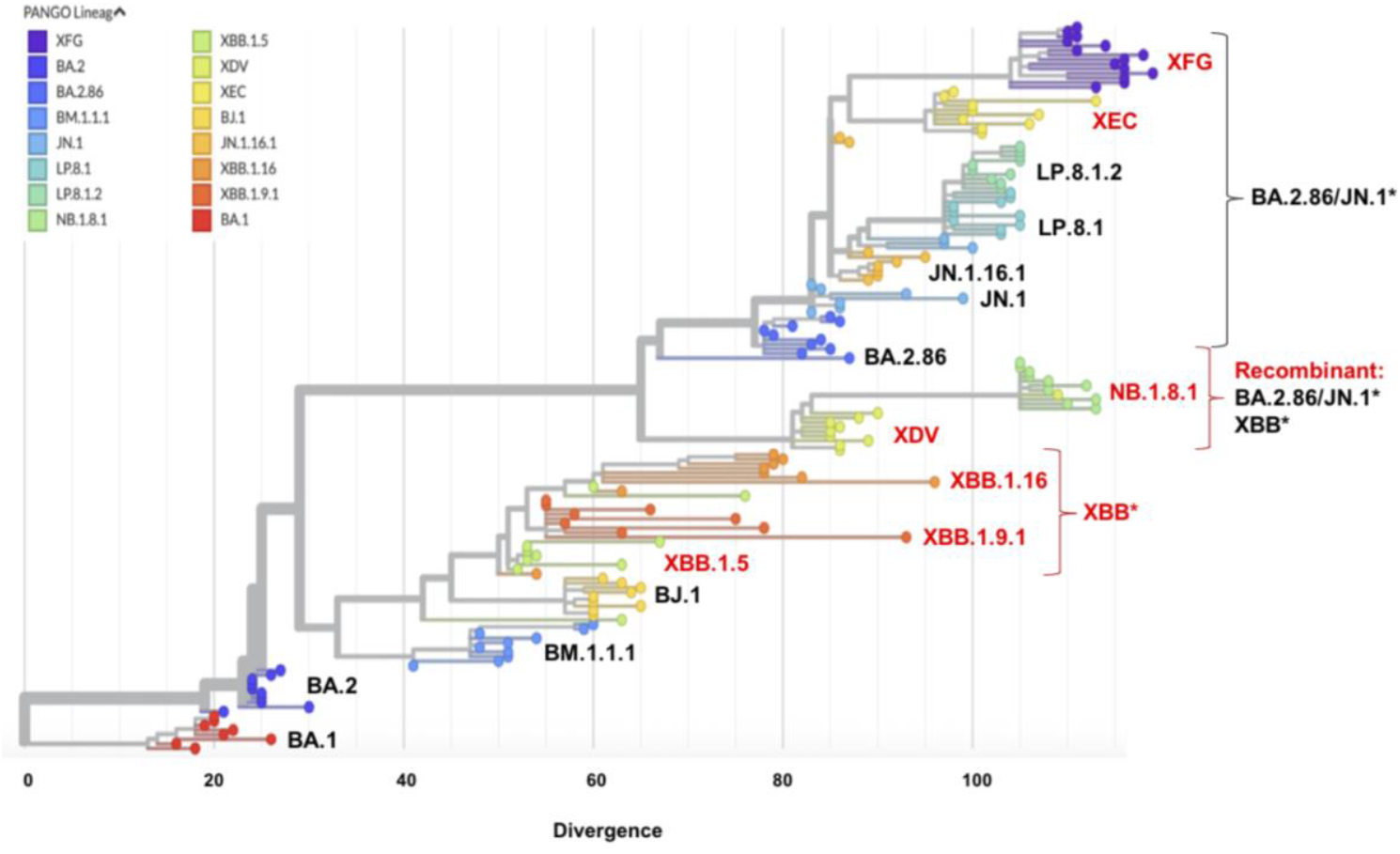
Recombination drives the evolution of SARS-CoV-2. Phylogenetic relationship between representative Omicron recombinant variants and the predicted original Omicron lineage is displayed. Each color represents one specific lineage. Numbers represent “Divergence” of each lineage from the predicted common ancestor of BA.1 and BA.2, the earliest Omicron lineages, at the position of divergence “0” (calculated through algorithm in Nextstrain). The names of the lineages are displayed next to the corresponding lineage groups, with recombinant variants being highlighted in red. The lineages belonging to the XBB* family or the BA.2.86/JN.1* family or recombinants of these two families are indicated. For each lineage, around ten randomly chosen samples are used in the analyses.

In addition to XBB.1.5, XBB.1.9.1 and XBB.1.16, XBB has produced many other subvariants, such as XDE, a recombinant variant of GW.5.1 (XBB.1.19.1.5.1) and FL.13.4 (XBB.1.9.1.13.4). In parallel to the circulation of XBB* lineage family, BA.2.86 and its subvariant JN.1 emerged and co-circulated with XBB* for more than 2 years (primarily in 2023 and 2024) ^26^, which has led to various recombination events between the XBB* lineage family and the BA.2.86/JN.1* lineage family. Among others, one product derived from recombination between these two big lineages is XDV, arising from recombination of JN.1 and XDE. NB.1.8.1 is a descendant lineage of XDV ^53,54^, which was identified as VUM and got spread worldwide ^25^. As shown in Figure 1, XDV and NB.1.8.1 are evolutionarily more closely related with the BA.2.86/JN.1* family than with the XBB* family, and NB.1.8.1 shows a higher divergence compared to the co-circulating JN.1 sub-lineages XEC, LP.8.1 and LP.8.1.2.

XEC was derived from recombination between two JN.1 sub-lineages: KS.1.1 (JN.1.13.1.1.1) and KP.3.3. XEC was also identified as VUM and got spread worldwide outcompeting many other JN.1 subvariants ^55,56^. LP.8.1 is another JN.1 subvariant that was identified as VUM ^25^. Interestingly, the divergence of both XEC and LP.8.1 is only slightly higher compared to JN.1. Similar to XEC, XFG is a recombinant variant arising from two JN.1 sub-lineages: LF.7 and LP.8.1.2, but XFG shows a much higher divergence compared to XEC, and also more evolved compared with NB.1.8.1 (Figure 1).

### 3.2 The spread and relative growth advantage of XFG in Germany

To know about the background for the spread of XFG in Germany, we first investigated the predominant SARS-CoV-2 variants in Germany in the spring and summer of 2025. Between January to March 2025, XEC and its sub-variants, labelled as XEC* in Figure 2A, were the predominant variants. Since April, with the emergence and spread of NB.1.8.1 and a few other variants, the frequency of XEC* reduced dramatically. In April, XFG was first detected in Germany. Although NB.1.8.1 and a few other lineages showed strong competitions, the spread of XFG outcompeted XEC, LP.8.1, NB.1.8.1 and other lineages. Till July 2025, the infection cases caused by XFG accounted for around 50% of the total cases (Figure 2A). The analyses were based on genome data from GISAID acquired on August 16, 2025.

**Figure 2:**
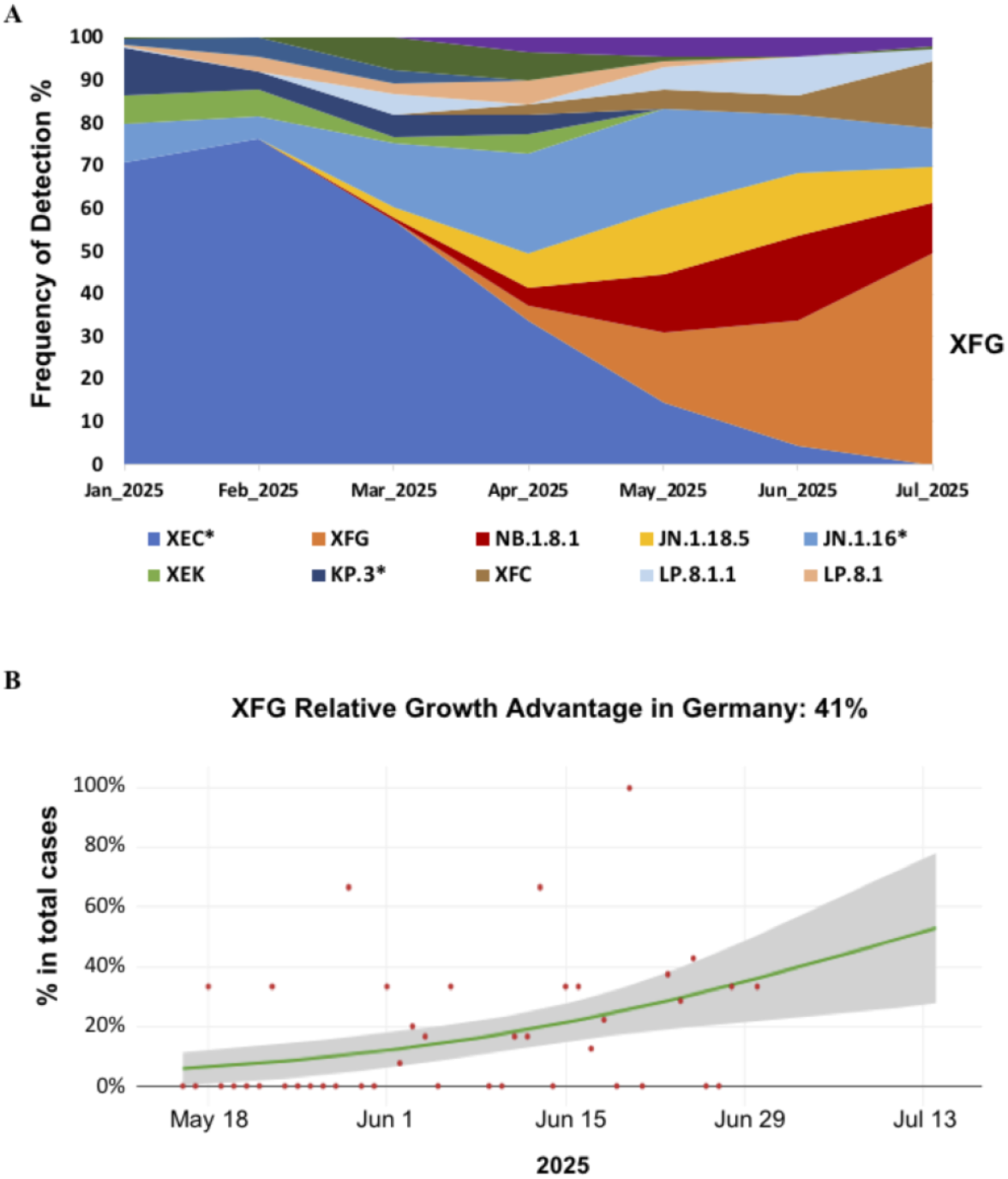
The emergence and spread of XFG in the spring and summer of 2025 in Germany and its relative growth advantage over co-circulating SARS-CoV-2 variants. **(A)** Lineage dynamic changes in Germany, January-July 2025. Frequency of detection (%) of each SARS-CoV-2 lineage in each month is displayed. To achieve a better resolution, if several sub-lineages of one variant were detected during this period, in most cases the sub-lineages are collectively shown together with the ancestor lineage, labelled as the ancestor lineage with star, such as XEC* indicating XEC and sub-lineages. **(B)** Growth of XFG during its early spreading period in Germany. Model fits are based on a logistic regression. Dots represent the daily proportions of variants. The relative growth advantage per week (in percentage; 0% means equal growth) is reported. The shaded areas correspond to the 95% CIs of the model estimates.

The fast spread of XFG suggests a relative growth advantage over co-circulating variants. Therefore, we analyzed the relative growth advantage of XFG during its early spreading period in Germany. The relative growth advantage reflects the advantage over co-circulating lineages in the selected region and time frame. As it has been revealed by previous studies that large scale community transmission often started or became detectable around one month later after the first sample was detected ^26^, to examine the growth advantage in the early spreading period, we focus on the time period of about 45 days right after community transmission started.

In Germany, during the early spreading period (between 2025-05-16 and 2025-06-30), XFG showed a relative growth advantage of 41% over co-circulating variants, mainly a few other JN.1 subvariants and NB.1.8.1 (Figure 2B). Considering NB.1.8.1 is one fast-spreading lineage itself with a 61% relative growth advantage globally in its early spreading period ^25^, the results indicate that XFG is characterized by a strong relative growth advantage over co-circulating variants.

## 4. Discussion

In recent several years, multiple SARS-CoV-2 lineages derived from recombination have become predominant variants worldwide, and the emergence of one widespread novel variant may usually cause one infection wave. In this study we evaluated the effects of recombination on SARS-CoV-2 evolution and investigated the growth of XFG in Germany as one example of the predominant recombinant variants. The information presented in this study indicates that SARS-CoV-2 genomic recombination between evolutionarily distant lineages or recently diverged lineages could both drive SARS-CoV-2 evolution. Furthermore, one clear relative growth advantage of XFG in Germany over co-circulating variants has been revealed, which might be partly attributed to the impact of recombination on the virus.

It has long been known that virus recombination is a common feature of sarbecovirus evolution ^57^. Studies indicated that before the establishment of SARS-CoV-2 in humans, recombination events among coronaviruses circulating in non-human species contributed to the evolution of coronaviruses ^7,8^. For other viruses, genomic recombination has been associated with evolution of HIV ^58^, human influenza viruses ^59^ and MERS-CoV ^60^. However, our understanding about the effects of recombination on SARS-CoV-2 evolution is still limited, and it remains unclear about the impact of genomic recombination on SARS-CoV-2 infectivity. The limited progress is partly owing to lack of large-scale epidemiological data, and partly owing to technical difficulties in charactering recombination lineages. In particular, it remains challenging to identify recombination between recently diverged lineages due to their highly similar sequence backgrounds. For such cases, mutations like insertions and deletions can be informative, especially owing to the fact that deletions usually could not revert during the evolution of a single lineage. Importantly, phylogenetic analyses may provide a tractable way to test hypotheses regarding virus recombination ^61^. However, the exact start and endpoints of recombinant genome regions, the so-called recombination breakpoints, must be inferred statistically for non-segmented viruses like SARS-CoV-2 ^62^. Moreover, it is also difficult to estimate the exact timing and location of recombination events owing to uncertainty in prediction of phylogenetic node ages, although such uncertainty can be minimized by integrating evolutionary information across different genome regions ^63^.

Owing to lack of epidemiological metadata, we can hardly perform large-scale analyses about the association between recombination events and public-health relevant parameters of SARS-CoV-2. The public genome databases built up through collective efforts of scientists worldwide, e.g. GISAID, make it possible to carry out genomic epidemiology investigation. As one example of the predominant recombinant variants, investigation about the growth of XFG has revealed that the emergence of XFG changed the SARS-CoV-2 spreading pattern in Germany. It has not only replaced XEC in the circulation, but also suppressed the growth of other co-circulating variants including LP.8.1 and NB.1.8.1. The widespread circulation of XFG should be at least partly on account of the effects of recombination. A few recent studies have compared the abilities of ACE2 binding and immune evasion among several prevalent co-circulating lineages, including LP.8.1, NB.1.8.1 and XFG. The studies indicate that XFG displayed robust antibody evasion stronger than LP.8.1 and NB.1.8.1 following either KP.2-adapted mRNA vaccine or LP.8.1-adapted mRNA vaccine ^64,65^. However, XFG has a relatively low ACE2 engagement efficiency ^64^, indicating lower receptor compatibility for sustained transmission. The fact that XFG displayed a relative growth advantage against other co-circulating variants, including LP.8.1 and NB.1.8.1, suggests that immune evasion might be a more decisive factor in virus spread, at least in the current status when there is established immune landscape in the population from previous infection or vaccination.

The fact that multiple recombinant variants, including XFG, have got spread worldwide and become predominant lineages in recent several years, indicates that certain recombination events may likely promote fitness, transmission/infection, or immune evasion ability of the virus by bringing unique mutation combination, which may possibly remodel the virus structure to certain extent or break certain structure limit facilitating novel mutations in the evolution course. In addition, recombination process may possibly clear deleterious mutations and overcome clonal interference ^10^. Through these possible approaches, after going through recombination, viruses may possibly acquire large “jumps” in certain functional aspect skipping many intermediate forms that might take a long time to acquire through cumulative mutations ^66-68^. It is therefore necessary to investigate further details about how recombination drives adaptation of SARS-CoV-2 for evaluating the long-term effects on virus evolution.

Furthermore, through the overview analysis regarding the evolution of the recombinant variants, the acquired information indicates that recombination between evolutionarily distant lineages (e.g. between XBB* and BA.2.86/JN.1*) or closely related variants (e.g. between two JN.1 subvariants) can both drive SARS-CoV-2 evolution, and has the potential of resulting in a novel predominant variant, e.g. NB.1.8.1 for the first scenario or XFG for the latter one. In addition to driving the evolution of SARS-CoV-2, genomic recombination also enriched the diversity of SARS-CoV-2 genetics. For example, XDV is one of the few designated tertiary recombinants (a recombinant of a recombinant of a recombinant), and the predominant variant NB.1.8.1 then emerged on the genetic background of XDV. These two variants display divergence from the XBB* family, and also show differences from the BA.2.86/JN.1* family, providing disparate directions for further evolution of SARS-CoV-2.

In summary, the results of this study revealed one important role of recombination in SARS-CoV-2 evolution, highlighting the importance of further investigating recombination events and tracking recombinant lineages, which is not only of significance for virus research but can also be critical for preventing future pandemic.

## Acknowledgements

We thank all researchers who generate and share genome data on GISAID (http://www.gisaid.org). We thank the Dresden-concept Genome Centre for their sequencing efforts.

## Data Availability

All data produced in the present work are contained in the manuscript.

## Code availability

Data processing and visualization was performed using publicly available software, primarily RStudio v1.3.1093. Code for constructing phylogenetic maximum likelihood (ML) and time trees is available at https://github.com/genomesurveillance/delta-variant-sublineage, which is modified from SARS-CoV-2-specific procedures github.com/nextstrain/ncov.

